# *Salmonella* Senftenberg isolated from wastewater is linked to a 2022 multistate outbreak

**DOI:** 10.1101/2024.02.20.24302949

**Authors:** Zoe S. Goldblum, Nkuchia M. M’ikanatha, Erin M. Nawrocki, Nicholas Cesari, Jasna Kovac, Edward G Dudley

**Affiliations:** Department of Food Science, The Pennsylvania State University, University Park, PA; The E. coli Reference Center, The Pennsylvania State University, University Park, PA; Pennsylvania Department of Health, Harrisburg, Pennsylvania, USA

**Author notes:** Corresponding author: Edward G. Dudley.

## Abstract

The ability of wastewater-based surveillance (WBS) to infer viral loads in communities was showcased during the COVID-19 pandemic. Whether WBS could be used to track other microorganisms is less studies. We tested for *Salmonella enterica*, a common foodborne pathogen, in community wastewater and investigated whether such isolates could be linked to past or ongoing outbreaks. During June 2022, composite wastewater samples were collected from treatment facilities in two communities in central Pennsylvania. A total of 42 confirmed *S. enterica* isolates were whole genome sequenced, revealing 7 different serovariants. Four isolates identified as *S. enterica* ser. Senftenberg were in a cluster, separated by 0-4 SNPs from clinical isolates uploaded to the web-based National Centers for Biotechnology Information Pathogen Detection platform. Most isolates in the cluster were previously associated with a 2022 multistate foodborne outbreak that was caused by contaminated peanut butter. Our targeted study suggests that wastewater-based surveillance for *Salmonella* would complement public health efforts by providing information about the extent of an outbreak and help focus public health resources and guidance to areas most impacted.

## Introduction

The ability of wastewater-based surveillance (WBS) to infer viral loads in communities was showcased during the COVID-19 pandemic.^1^ A few studies have also demonstrated that bacterial pathogens such as *Salmonella enterica* can be simultaneously isolated from wastewater and clinical cases.^2, 3, 4^ It is unclear to what extent routine surveillance for bacterial pathogens would benefit public health. To evaluate this, we screened samples from two wastewater facilities for *S. enterica* during June 2022, concurrent with a study investigating SARS-CoV-2 presence. We additionally asked whether we could isolate *S. enterica* that matched human isolates from ongoing outbreaks.

## Methods

Composite wastewater samples were collected twice per week during June 2022, from two wastewater treatment facilities in central Pennsylvania, designated WWTP-1 and WWTP-2 (Table 1). These facilities treat wastewater from populations of approximately 3,600 and 13,600 individuals, respectively. *S. enterica* isolates were identified by adapting protocols from the FDA Bacteriological Analytical Manual. Briefly, 120 mL of wastewater was centrifuged at 5,000 *g* for 20 min at 4°C to precipitate solids. The supernatant was passed through a 0.45 μm filter, and the filter was added to the pellet along with 40 mL buffered peptone water supplemented with 20 mg/L novobiocin. After 24h recovery at 35°C, the samples were subcultured into selective media (Tetrathionate and Rappaport-Vassiliadis broths) and incubated at 42°C for 24h. Aliquots of 10 μl of each enrichment culture were plated on Xylose Lysine Deoxycholate and Hektoen enteric agars and monitored for growth and colonies characteristic of *Salmonella*. Putative *S. enterica* colonies were restreaked to purify and identity was confirmed by PCR targeting *invA*. Libraries were constructed using the Nextera XT DNA library kit and sequenced on an Illumina MiSeq using the MiSeq reagent v3 kit with 500 (2 × 250) cycles. Short sequence reads were uploaded to the National Center for Biotechnology Information Pathogen Detection site (https://www.ncbi.nlm.nih.gov/pathogens) for genomic comparison to human clinical isolates and identification of molecular serovars. An annotated phylogenetic tree was constructed by importing the Newick data from Pathogen Detection into the Interactive Tree of Life (https://itol.embl.de/). Cluster verification was done on the CDC’s System for Enteric Disease Response, Investigation, and Coordination (SEDRIC).

**Table 1.** *S*. Senftenberg isolated from Pennsylvania wastewater treatment facilities.

| **Isolate** | **Isolation date** | **Location** | **Biosample accession** |
| --- | --- | --- | --- |
| PSU-5375 | June 15, 2022 | WWTP-1 | SAMN33902330 |
| PSU-5376 | June 15, 2022 | WWTP-1 | SAMN33902331 |
| PSU-5387 | June 22, 2022 | WWTP-1 | SAMN33902342 |
| PSU-5398 | June 16, 2022 | WWTP-2 | SAMN34154796 |

## Results

A total of 42 *Salmonella* isolates were obtained from wastewater samples collected between June 13th and June 29th, 2022. Whole genomes were sequenced, and isolates were identified as serovars Panama (38.1%), Senftenberg (21.4%), Baildon (19.0%), Agona (7.1%), Oranienburg (7.1%), Montevideo (4.8%) and Kintambo (2.4%). Four ser. Senftenberg isolates were separated by 0-4 single nucleotide polymorphisms (SNPs) from a cluster of 40 clinical isolates uploaded to Pathogen Detection between March 2022 and April 2023 (**Table 1, Fig. 1**). Using SEDRIC we verified that 21 (52.5%) of the clinical isolates matched a 2022 multistate foodborne outbreak (designated cluster 2205MLJMP-1 in SEDRIC) that was caused by contaminated peanut butter. Patients lived in 17 different states.^5^ While Pennsylvania was not one of these, three additional Senftenberg matching 2205MLJMP-1 were isolated from three patients in Pennsylvania after the outbreak investigation ended on May 9th, 2022.

**Figure 1:**
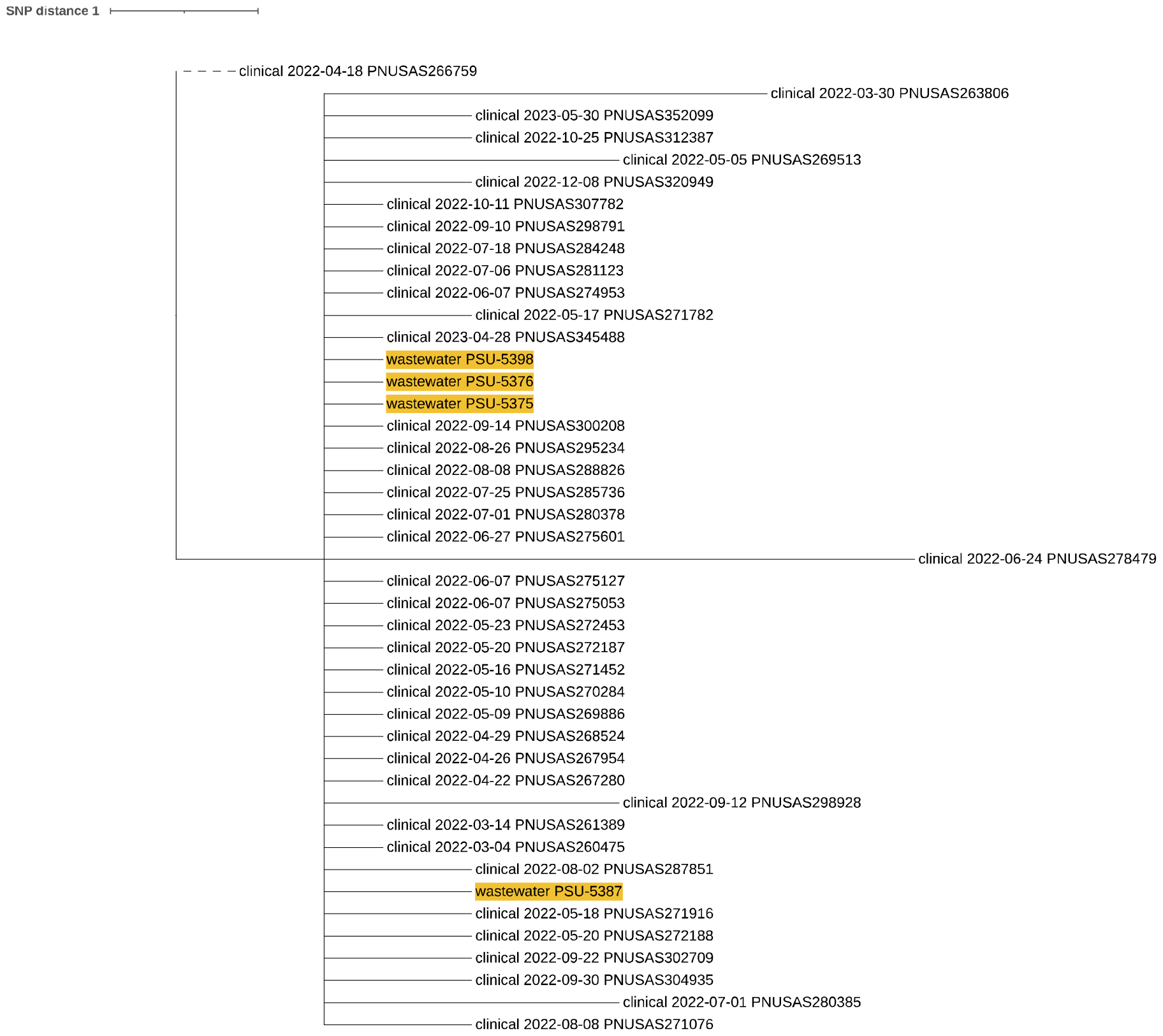
Wastewater isolates of S. Senftenberg are genetically linked to those associated with a 2022 multistate outbreak. SNP-based tree constructed using Newick data generated by the NCBI Pathogen Detection website, showing the relationship between the four *S*. Senftenberg isolates reported here (in yellow) and previously whole genome sequenced human isolates within the same cluster.

## Discussion

From two rural wastewater treatment facilities, we isolated *S*. Senftenberg that were genomic matches to strains associated with a multistate outbreak. No human cases were reported in Pennsylvania, which may be due to under-reporting of non-Typhoidal *Salmonella*.^6^ Our targeted study suggests that wastewater-based surveillance for *Salmonella* would complement public health efforts by providing information about the extent of an outbreak, and help focus public health resources and guidance to areas most impacted. It may also help identify where unreported cases reside, assist with targeted prevention messaging, and with public health actions including recall audit checks.

## Data Availability

All sequencing data was deposited in the NCBI Sequence Read Archive (SRA) (https://www.ncbi.nlm.nih.gov/sra) under BioProject PRJNA357723.

https://www.ncbi.nlm.nih.gov/bioproject/?term=PRJNA357723

## Obtained funding

Dudley, M’ikanatha, and Kovac

## Administrative, technical, or material support

Goldblum, Nawrocki, Cesari

## Supervision

Dudley, M’ikanatha, Kovac

## Conflict of Interest Disclosur

Dr. Dudley has received grants with salary support during the conduct of this study from the US Food and Drug Administration.

## Data Access, Responsibility, and Analysis

Dr. Dudley had full access to all the data in the study and takes responsibility for the integrity of the data and the accuracy of the data analysis.

## Acknowledgements

We thank Drs. Shannon McGinnis (Pennsylvania Department of Health) and Steven Ostroff (Ostroff Consulting) for reviewing this letter prior to submission. We thank the CDC’s System for Enteric Disease Response, Investigation, and Coordination (SEDRIC) for permission to conduct verification of isolates from NCBI and Michael Vasser with the CDC’s Foodborne Outbreak Response Team for insights on genomic analysis.

## References

1. The potential of wastewater-based epidemiology. Nat Water 2023; 1 (399). doi: 10.1038/s44221-023-00093-6

2. Diemert S, Yan T. Clinically unreported Salmonellosis outbreak detected via comparative genomic analysis of municipal wastewater Salmonella isolates. Appl Environ Microbiol. 2019;85(10):e00139–19. doi: 10.1128/AEM.00139-19.

3. Kuhn, KG, Shukla, R., Mannell, et al. Using wastewater surveillance to monitor gastrointestinal pathogen infections in the State of Oklahoma. Microorganisms. 2023; 11:2193. doi: 10.3390/microorganisms11092193

4. Sahlström L, de Jong B, Aspan A. Salmonella isolated in sewage sludge traced back to human cases of salmonellosis. Lett Appl Microbiol. 2006; 43(1):46–52. doi: 10.1111/j.1472-765X.2006.01911.x.

5. United States Centers for Disease Control. Where sick people lived. https://www.cdc.gov/salmonella/senftenberg-05-22/map.html. Accessed January 28, 2024.

6. Scallan E, Hoekstra RM, Angulo FJ, et al. Foodborne illness acquired in the United States--major pathogens. Emerg Infect Dis. 2011;17(1):7–15. doi:10.3201/eid1701.p11101.

